# Gabapentin Treatment Patterns Among Older Patients After Hospital Discharge for Acute Ischemic Stroke

**DOI:** 10.1101/2025.09.23.25336477

**Authors:** Shuo Sun, Maria A. Donahue, Rebeka Bustamante Rocha, Madhav Sankaranarayanan, Joseph P. Newhouse, Sonia Hernandez-Diaz, Alexander C. Tsai, Sebastien Haneuse, Lidia M.V.R. Moura

## Abstract

**Importance:** Gabapentin is frequently prescribed off-label for pain management following an acute ischemic stroke. However, little is known about optimal gabapentin prescribing practices, such as appropriate treatment duration for specific indications, particularly off-label uses.

**Objective:** We examined gabapentin treatment patterns among older adults who initiated gabapentin within 30 days of discharge from hospitalization for acute ischemic stroke.

**Design:** This is an observational, retrospective study of existing administrative claims data of Medicare beneficiaries with acute ischemic stroke (AIS) hospitalizations.

**Setting:** A national 20% random sample of US Medicare beneficiaries.

**Participants:** Patients aged 65 years and older who were hospitalized for their first acute ischemic stroke between 2013 and 2021.

**Exposures:** Gabapentin initiation within 30 days of discharge.

**Main Outcomes and Measures:** A novel approach that combines a time-varying proportion of days covered, calculated every two months, with a latent class mixed model to identify and characterize gabapentin treatment patterns during the first 12 months after initiation, accounting for prescription overlap and hospitalizations. An 80% proportion of days covered threshold was applied within each interval to distinguish high versus low medication coverage.

**Results:** The analytic cohort (N=1,628) had a mean age of 76.4 (IQR 70-82) years and was 60% female and 76.5% non-Hispanic White. The latent class mixed model identified three distinct gabapentin treatment patterns: 692 patients (42.5%) experienced rapid low medication coverage (proportion of days covered <80%) within two months after initiation; 96 patients (5.9%) experienced gradually declining medication coverage over 8 months after initiation; and 840 patients (51.6%) maintained high and stable medication coverage for at least one year.

**Conclusions and Relevance:** In this nationwide sample, half of older adults hospitalized for acute ischemic stroke and who initiated gabapentin within 30 days of discharge had gabapentin coverage for 12 months or longer after initiation.

**Key Points:** *Question:* Are stroke survivors aged 65 and older receiving a gabapentin prescription for longer than 12 months?

*Findings:* after analyzing a 20% sample of Medicare beneficiaries, 51.6% of older adults hospitalized for an acute ischemic stroke initiated gabapentin within 30 days of discharge and had gabapentin coverage for 12 months or longer after initiation.

*Meaning:* Most stroke survivors aged 65 and older continue receiving a gabapentin prescription 12 months after their stroke.

## INTRODUCTION

Approximately 795,000 strokes occur in the US every year.^1^ Acute ischemic stroke (AIS) is the most common type, and it can lead to several complications, including seizures, dysphagia, anxiety, depression, and post-stroke pain.^2,3^ Gabapentin is an antiseizure medication (ASM) used off-label to manage post-AIS pain,^4–6^ but its use raises concern given medication side effects and AIS complications, particularly among older age persons who often present with multiple comorbidities and high rates of polypharmacy.^7^ Despite these concerns, the prevalence of gabapentin use among older adults more than tripled from 2.6% in 2002 to 8.4% in 2015,^8,9^ and further increased to 9% in 2019-2021.^10^ Up to 95% of prescriptions are estimated to be given for off-label indications related to chronic pain,^11^ such as neuropathic, postoperative, and malignancy-related pain.^4,12,13^

While gabapentin treatment duration post-AIS varies case-by-case, based on patient response and tolerability, animal studies have demonstrated motor function improvement with daily gabapentin treatment for six weeks post-stroke.^14,15^ However, there are no clear, universal guidelines for the use of gabapentin after AIS.^5,14,15^ Limited safety and effectiveness studies examining its long-term use raise serious safety concerns and adverse effects that have been linked to gabapentin use, including sedation, respiratory depression, heart failure, and risk of misuse, especially among older adults who are already at risk due to polypharmacy, drug-drug interactions, impaired drug metabolism, and comorbidities.^11,16–20^ Although initiation of gabapentin in multiple conditions is well-documented, there is weak and inconsistent literature on its proven effectiveness and post-initiation use for post-stroke pain, and treatment patterns remain poorly understood.^10,11,21^

We examined gabapentin treatment patterns over the first 12 months following initial prescription. We used a measure of medication coverage, proportion of days covered (PDC), quantifying the percentage of days a patient has access to the medication during a given period.^22,23^ In this study, we defined three treatment patterns or “classes” based on patients’ medication coverage trajectories, calculated as PDC every two months during the 12 months post-initiation. We defined high and low medication coverage over time using the 80% threshold endorsed by the Pharmacy Quality Alliance (PQA) and the US CDC.^24^ Then we applied latent class trajectory analysis to the two-month interval-specific PDCs to identify distinct gabapentin treatment patterns.

## METHODS

This study followed the Strengthening the Reporting of Observational Studies in Epidemiology (STROBE; Table S1)^25^ reporting guidelines and was approved by the Mass General Brigham Institutional Review Board. Because we conducted a secondary analysis of data collected for routine billing purposes, the requirement for informed consent was waived. The data supporting this study’s findings were collected by the US Centers for Medicare & Medicaid Services (CMS) and were made available by CMS with no direct identifiers. All results were aggregated following the CMS Cell Size Suppression Policy. Restrictions apply to the availability of these data, which were used under license for this study. Medicare data are available through CMS with their permission.

### Study Cohort

Using national administrative claims data, we analyzed a 20% sample of U.S. Medicare beneficiaries aged 65 and older hospitalized for AIS between April 1, 2013, and September 30, 2021, who initiated gabapentin in an outpatient setting within 30 days of discharge, typically when patients would get their first outpatient refill.^26^

We included each patient’s first hospitalization for AIS observed in the 2014-2020 data window (hereafter referred to as the “index admission”), and applied a one-year lookback period to assess recurrent AIS.^27–29^ For each patient, time 0 was defined as the date of the first gabapentin prescription filled in Part D within 30 days after discharge from the index admission. To ensure sufficient observation time after the index admission, eligible patients had to either have at least one year of enrollment in prescription drug coverage (“Part D”) after initiation or die within one year of initiation post-discharge.

### Data Source

Hospitalization information was extracted from the Medicare Provider Analysis and Review (MedPAR) database based on principal diagnosis codes for AIS. We used International Classification of Diseases, 9th Revision (ICD-9) codes 433, 434, and 436, and ICD-10 codes I63.x,^30^ a validated strategy to capture acute stroke in administrative databases.^31^ Demographic variables, including age, sex, race, ethnicity, insurance status, and mortality, were obtained during AIS diagnosis for all study patients from the Medicare’s Master Beneficiary Summary (MBSF) File. Patient characteristics such as seizure diagnosis, pain, and depression history were included to improve the accuracy and clinical relevance of subgroup classification.

We included beneficiaries who were enrolled in traditional fee-for-service Medicare Parts A (inpatient and other services), B (outpatient), and D (prescription drug), continuously for 12 months before their AIS admission, as described in more detail below. Data from beneficiaries enrolled in Part C Medicare Advantage, a potentially more affordable managed care model alternative to traditional Medicare coverage, were unavailable for this study.^32^

## Measuring Gabapentin Treatment Patterns

We analyzed prescription claims data to measure gabapentin treatment patterns in the first 12-month period following initiation. Outpatient prescription claims are available in the Medicare Part D Event File.^33^ Prescription claims include records of when an individual fills an outpatient medication from the pharmacy and is billed through insurance. The claims include the date the prescription was filled, medication name, National Drug Code (NDC), quantity dispensed, and days’ supply, as well as other variables for new prescriptions and refills. Part D data does not include medication indication or inpatient prescriptions; these are billed as part of hospital stays.

The standard PDC metric calculates the proportion of days within a defined period of 12 months that a patient has access to their medication, considering the number of days available for refills to cover the medication.^34,35^ PDCs are widely used in claims-based medication coverage research.^22,23^ Previous studies have used claims data to calculate PDCs to measure coverage to various drug classes.^36–38^ In this study, we used a series of pragmatic strategies to construct an adjusted PDC measure. Specifically, inpatient hospitalizations were classified as either short or long stays (criteria detailed below), with subsequent PDC values set to zero after extended stays; overlapping prescriptions were adjusted by carrying forward Surplus days, and follow-up was truncated at death. When a patient refilled gabapentin before the previous supply was finished, overlapping days were carried forward (“stockpiled”) so that surplus doses extended subsequent coverage rather than inflating PDC values. The same adjustment was applied to any surplus doses dispensed during hospitalization. The numerator of PDC represented the number of days covered by gabapentin after these adjustments. The denominator was defined over 60-day intervals, censored at the date of death when applicable. Figure 1 illustrates an example of these strategies for constructing the adjusted PDC measure.

**Figure 1.**
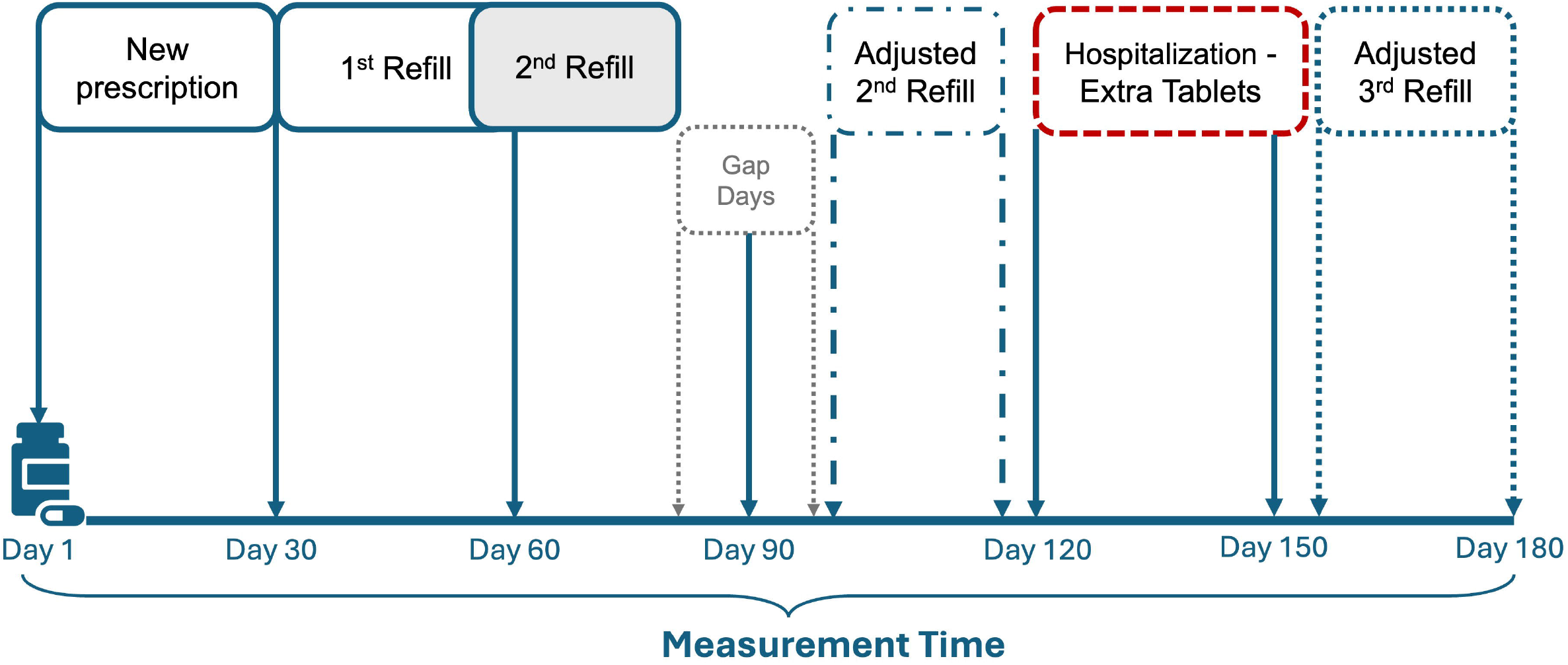
Hospitalization and Overlap-Adjusted PDC. An example of the hospitalization and overlap-adjusted Proportion of Days Covered (PDC). ^*^For non-disruptive hospitalization. In this example, the patient initiated gabapentin within 30 days post-discharge and experienced two coverage gaps after the first and second refill periods. The patient then had an inpatient hospitalization (in this example, lasting less than 14 days). Because the patient received gabapentin from the hospital pharmacy during the inpatient admission, they had extra doses of gabapentin remaining at home. The date of the third refill was therefore shifted forward.

To construct the trajectories, we divided the 12-month follow-up period into consecutive 60-day intervals (since the average prescription days’ supply is 30)^26^. We calculated the adjusted PDC for each patient within each interval, resulting in seven observations per patient. An 80% threshold was used to define high or low medication coverage.

As mentioned above, inpatient medication claims are not captured in Part D; charges for the medication are bundled with other inpatient services and do not appear separately. We adjusted hospital stays after the initial gabapentin prescription based on hospitalization length to avoid underestimating medication coverage:^39^

1. Hospitalization lasting 14 days or less: We assumed patients had inpatient gabapentin coverage during hospitalization. When discharged, they would likely have extra pills. This could make it seem as if they were late refilling their prescription, which would underestimate their gabapentin PDC. To account for this, we assumed these patients had a surplus of medication equivalent to the length of stay.
2. Hospitalizations lasting more than 14 days: These were not considered disruptions if patients refilled their gabapentin prescriptions within 90 days after discharge from these hospitalizations. Patients were assumed to have an extra 14 days’ worth of medication doses, and their medication supply was adjusted to ensure accurate calculations. If patients did not refill their prescription within 90 days of discharge, the days following the hospital admission, their subsequent PDC values were set to zero.

### Statistical Analysis

To address the limitation of using a single summary measure of gabapentin use, we leveraged trajectory modeling to capture time-varying patterns. This approach allowed us to detect behaviors such as prescription inertia (treatment coverage declining to near zero rapidly), progressive tapering (gradual low coverage across time), and sustained use (stable high coverage). We applied a latent class mixed model^40^ to the adjusted PDC data trajectories, adjusting for prescription overlaps, hospitalization, and death (as in Measuring Gabapentin Treatment Patterns Section), to identify and characterize gabapentin treatment patterns during the first 12 months following initiation.

The latent class mixed model conceptualizes the patient population as a mixture of G unobserved subgroups, each characterized by a unique mean trajectory. These G mean trajectories are constructed as a function of the time variable and covariates using latent class-specific mixed effects models. The standard linear mixed model assumes that subjects are from a homogeneous population and estimates one mean trajectory, accounting for subject-level variation with random effects. In contrast, the latent class mixed model permits additional flexibility by allowing for a heterogeneous population with latent sub-groups, each characterized by its distinct mean trajectory profile. This approach is beneficial when different subgroups follow different medication patterns over time. Specifically, each individual is assumed to belong exclusively to a single latent class, represented by a discrete random variable *c*_*i*_ which takes the value g if the individual *i* is classified *i* into class g (g = 1,…,G).^41^ Since *c* is unobserved, its distribution is modeled using a multinomial logistic regression with covariates. For the outcome, we applied a logistic regression to a binary variable coded as 1 for low coverage periods and 0 otherwise. Model selection was guided by the Bayesian Information Criterion (BIC), and all analyses were carried out using the lcmm package in R (version 4.4.0).

## RESULTS

### Sample Characteristics

Our sample included 1,628 patients (Figure S1). The median age was 76 years (interquartile range [IQR], 70-82); 60% were women and 80% were White (Table 1). About half (820) were admitted to an inpatient setting during the year of observation, and 15% died. Among the 820 patients with at least one admission within the study period, 82% had a length of stay less than or equal to 14 days (median, 5 days; IQR, 3-12), and 742 had at least one refill after discharge from hospitalizations. Most patients in the sample had gabapentin coverage for 12 months after initiation, followed by those with shorter prescription coverage of less than two months.

**Table 1.**
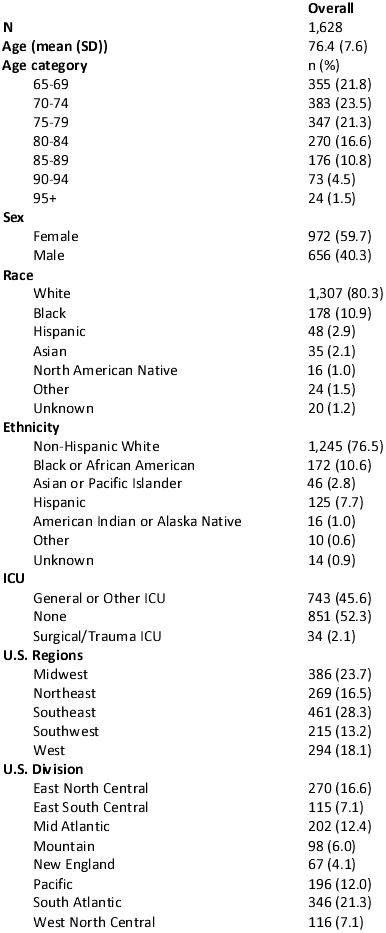
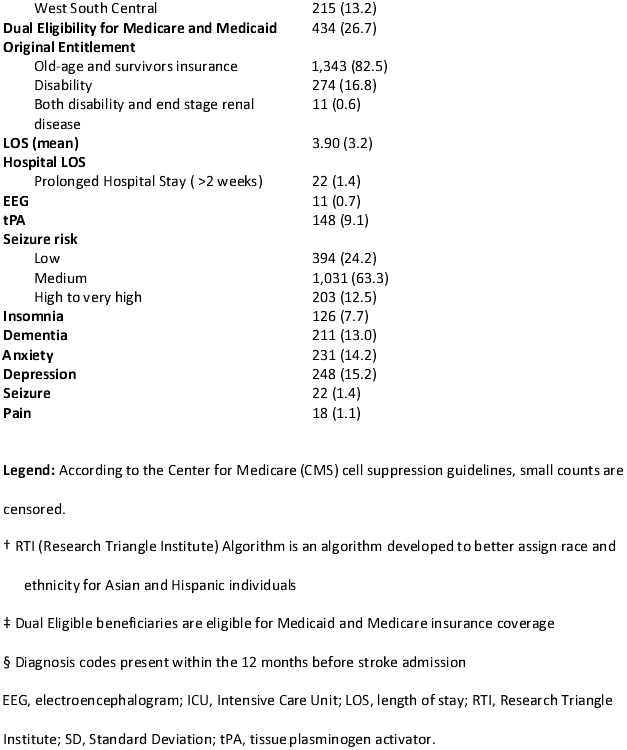
Sample Characteristics.

### Latent Class Mixed Model

We fitted models with 1 to 4 classes under various specifications (Table S2). The optimal model yielded three classes with a BIC of 9,054 (Table S2). We illustrate the classification of gabapentin treatment patterns in the optimal model in Table 2. We summarized the estimated parameters, standard errors, Wald test statistics, and the corresponding p-values of the optimal latent class mixed model in Table S3. We did not calculate the class 1 intercept in the longitudinal; it’s fixed to serve as the reference group to ensure the model is statistically identifiable.^40^

**Table 2.** Patient characteristics by gabapentin treatment patterns identified from the optimal model.

|  | **Class I** | **Class II** | **Class III** |
| --- | --- | --- | --- |
| Number of patients (%) | 692 (42.5%) | 96 (5.9) | 840 (51.6) |
| Age, mean (SD) | 76.4 (7.5) | 75.6 (6.8) | 76.6 (7.8) |
| Female, N (%) | 396 (57.2) | 56 (58.3) | 520 (61.9) |
| Baseline Seizure, N (%) | < 11(*) | < 11(*) | < 11(*) |
| Baseline Pain, N (%) | < 11(*) | < 11(*) | < 11(*) |

**Legend:** Class I (rapid lower medication coverage)- those who had <80% PDC of gabapentin within the first 2 months after initiation. Class II (gradual declining coverage)- those demonstrating a declining PDC (from 80% to 0%) around 8 months post-initiation. Class III (high and stable medication coverage) - patients with adjusted PDC values >80% for at least 12 months after initiation. \*Counts and percentages have been suppressed following the NIH Cell Suppression Policy.

The optimal model identified the treatment patterns as follows: 42.5% for Class I: early and rapid low medication coverage (PDC <80%) within first 2 months, 5.9% for Class II: gradual declining coverage (PDC <80% from the second month to eighth month after initiation), and 51.6% for Class III: stable high coverage (PDC ≥ 80%). Table 2 summarizes the characteristics of these three classes, including demographic variables, baseline seizure, pain, and depression. The continuous PDC trajectories stratified by the three classes are shown in Figure S2.

Patients with baseline seizure events were more likely to fall in Class I or II (*p*-value0). Age, sex, baseline pain, and depression were not significantly associated with Class I or II. We plotted the comparison between females aged 75 without depression or pain, stratified by the presence or absence of a baseline seizure (Figure 2). Figure 2 visually illustrates the three classes from Table 2 through distinct patterns in predicted low gabapentin coverage probability curves, stratified by baseline seizure status. Class I curves increase sharply after the first 60 days, reflecting early and rapid low coverage to treatment. Class II curves rise gradually, with low coverage probabilities exceeding 0.5 around month 8. Class III shows flat curves with low probabilities, indicating consistently high medication coverage. Within each class, patients with baseline seizures tended to have higher predicted low medication coverage probabilities than those without seizures, suggesting shorter gabapentin coverage.

**Figure 2.**
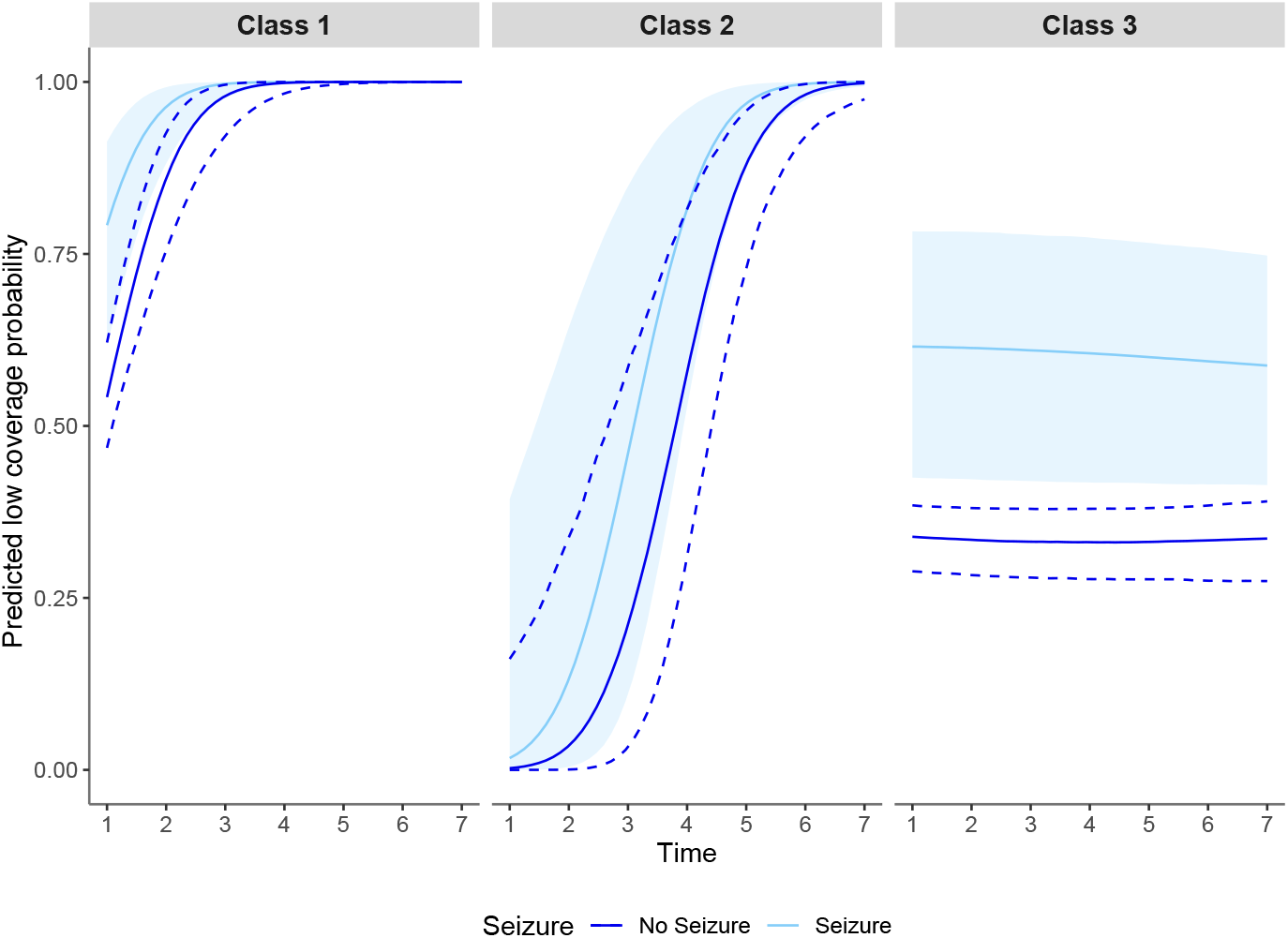
Comparison of females aged 75 without depression or pain, by baseline seizure status. **Legend:** Class I (rapid lower medication coverage)-those who had <80% PDC of gabapentin within the first 2 months after initiation. Class II (gradual declining coverage)-those demonstrating a declining PDC (from 80% to 0%) around 8 months post-initiation. Class III (high and stable medication coverage) - patients with adjusted PDC values >80% for at least 12 months after initiation. Dark blue: no seizure; light blue: seizure.

## DISCUSSION

Using a national sample of Medicare beneficiaries and stroke survivors aged 65 years and older, we identified three distinct gabapentin treatment patterns. We used time-varying PDCs with explicit assumptions regarding prescription overlap, inpatient stays, excess medication, and death as a censoring factor. Additionally, we applied a latent class model adjusting for relevant clinical covariates to ensure that the classes reflected distinct treatment patterns and to allow exploration of their association with patient characteristics. This nuanced approach, supported by a transparent analytical framework, can be replicated in future research on other medications. Lastly, standard implementations often overlook factors such as overlapping prescriptions and hospitalizations.^34,42,43^

Gabapentin was initially approved by the FDA in 1993 as an antiseizure medication (ASM), and later in 2002, received an indication for post-herpetic neuralgia.^44^ Despite limited evidence on its effectiveness, or its long-term safety in older adults, and existing guidelines that recommend against prolonged use in this population, more than half of patients remained on gabapentin treatment for the 12-month follow-up.^45^ Most of these patients did not have indications (e.g., seizures) or off-label indications (e.g., pain, insomnia, dementia, anxiety, depression).

Notably, FDA approval for gabapentin is limited to adjunct treatment of partial-onset seizures, postherpetic neuralgia, and restless legs syndrome (gabapentin enacarbil).^44^ Gabapentin may also be considered off-label for treating central post-stroke pain. However, its efficacy in this context is not well-studied but is based on its use in the treatment of other forms of neuropathic pain.^46,47^ The Guidelines for Adult Stroke Rehabilitation and Recovery recommend gabapentin as second-line therapy, along with other ASMs such as pregabalin, carbamazepine, and phenytoin.^48^ Because of its use in managing chronic pain syndromes, gabapentin is frequently prescribed concomitant with opiates, semi-synthetic opioids, synthetic opioids, and/or benzodiazepines, with 10-30% of older adults reporting concomitant use.^49^ Such polypharmacy is explicitly discouraged by both the FDA and the American Geriatrics Society, given the increased risk of severe sedation-related adverse events, such as respiratory depression and death.^45^

Even though most patients in our sample did not have a history of seizures at baseline, those who did were categorized as Class I or II (rapid and gradual declining low coverage). However, 63% of the sample were at medium risk for seizures (Table 1), although prophylactic use of ASMs after stroke is not recommended.^14,45,49–51^ For patients with recurrent seizures, medication should be tailored based on individual characteristics, and gabapentinoids should be used judiciously,^52^ especially among older adults who are particularly vulnerable to gabapentinoid side effects (e.g., hip fractures, edema, congestive heart failure, dementia, breathing difficulties, and atrial fibrillation),^16,18–20^ central nervous system depression,^49,53–55^ and renal function decline.^12,20^

Incidentally, we found that in our sample, nearly half of the patients were admitted to an inpatient setting within 12 months following gabapentin initiation. While we cannot determine causation, this finding raises concerns and underscores the need for further studies and closer monitoring of older adults prescribed these drugs. The complexity of gabapentin treatment patterns is influenced by perceived efficacy, out-of-pocket cost,^56^ forgetfulness, drug interactions (given the high rates of polypharmacy), and side effects.^18,20,52,57^

Since gabapentin is widely used off-label,^58^ a lack of clear guidelines on optimal dosage and treatment duration may contribute to chronic prescribing.^49,57^ Treatment patterns in older adults are complex and influenced by several factors, including perceived efficacy, out-of-pocket cost,^56^ forgetfulness, drug interactions (given the high rates of polypharmacy), and side effects.^18,20,52,57^ Our study is the first to provide insights into real-world gabapentin treatment patterns among older Medicare stroke survivors.

### Limitations

Our findings may not be generalizable to younger populations or the Medicare population in Part C (Medicare Advantage), a little over half of the US Medicare population. Medicare Advantage enrollees continue to increase over time, and studies have reported that vulnerable groups are more likely to enroll in Medicare Advantage than traditional fee-for-service Medicare.^32^ They also do not include patients discharged to skilled nursing facilities or inpatient rehabilitation units, because we focused on outpatient medication use and treatment patterns.

Although claims-based data are generally reliable, they have limitations, including entry errors and missing baseline variables. We computed PDC using the recorded prescription duration (days’ supply). Because we did not account for post-fill dose changes or drug switches, PDC may reflect dispensations and patients’ coverage for a specific medication. Still, it does not reflect actual consumption, potentially underestimating when doses increase and overestimating when doses decrease or overlapping supplies occur. Additionally, dichotomizing PDCs into high and low coverages would result in a loss of information.

### Future Directions

Few large-scale, inclusive studies have characterized gabapentinoid treatment patterns and their impact on older adults.^12,20,53,57^ Future studies could explore the safety and effectiveness of long-term gabapentin use (i.e., longer than six and 12 months), side effects, differences in pain expression, pain management strategies, care quality, and outcomes, among various vulnerable populations, such as patients with Alzheimer’s and Related Dementias, cardiovascular disease, and cancer.

## CONCLUSION

Notably, most older adults who were hospitalized for AIS and who initiated gabapentin within 30 days of discharge had gabapentin coverage for 12 months or longer. This highlights the need for clear prescription guidelines and more studies on the long-term safety and effectiveness of gabapentinoid use among older adults.

## DATA AVAILABILITY

We had a Data Use Agreement approved by the Center for Medicare & Medicaid Services (DUA RSCH-2022-58182). Interested researchers may replicate the study by obtaining the data from CMS. Reproducing this study requires 20% MedPAR, Outpatient, Carrier, and Part D standard analytical files.

## ACKNOWLEDGEMENTS

S.S. completed the data analysis, drafted the methods and results sections, and revised the manuscript for intellectual content. M.A.D. drafted, edited, and revised the manuscript for intellectual content. R.B.R. edited and revised the manuscript for intellectual content. J.P.N. facilitated data access and revised the manuscript for intellectual content. A.T., S.H.D., and S.H. were responsible for study design, conceptualization, supervising the analysis, and revising the manuscript. L.M.V.R.M. was responsible for study design, conceptualization, facilitating data access, supervising the analysis, drafting, and revising the manuscript.

## SOURCES OF FUNDING

This study was supported by the NIH (1R01AG073410-01).

## DISCLOSURES

S.S., M.A.D., R.B.R., S.H.D., and S.H. have no conflict of interest to disclose.

J.P.N. is the National Committee for Quality Assurance director and reports no conflict of interest. A.C.T. has received support from the NIH and reports receiving financial honoraria from Elsevier (for his role as Co-Editor in Chief of the Elsevier-owned journal *SSM – Mental Health*) and BMJ Publishing Group Ltd. (for his role as Clinical Editorial Advisor of the BMJ Publishing Group Ltd.-owned journal *The BMJ*). He acknowledges salary support from NIH K24DA061696-01.

L.M.V.R.M. receives support from the NIH, CDC, and Epilepsy Foundation, and reports no conflict of interest.

## Notes

### Competing Interest Statement

The authors have declared no competing interest.

### Author Declarations

Ethics committee/IRB of Mass General Brigham Institutional Review Board gave ethical approval for this work.

