## Supplemental Figures 1, 2, Tables 1, 2, 3, and code for "Gabapentin Treatment Patterns Among Older Patients After Hospital Discharge for Acute Ischemic Stroke"

**Figure S1. PRISM Diagram Acute Ischemic Stroke Cohort Flow Diagram, 20% Medicare Data**

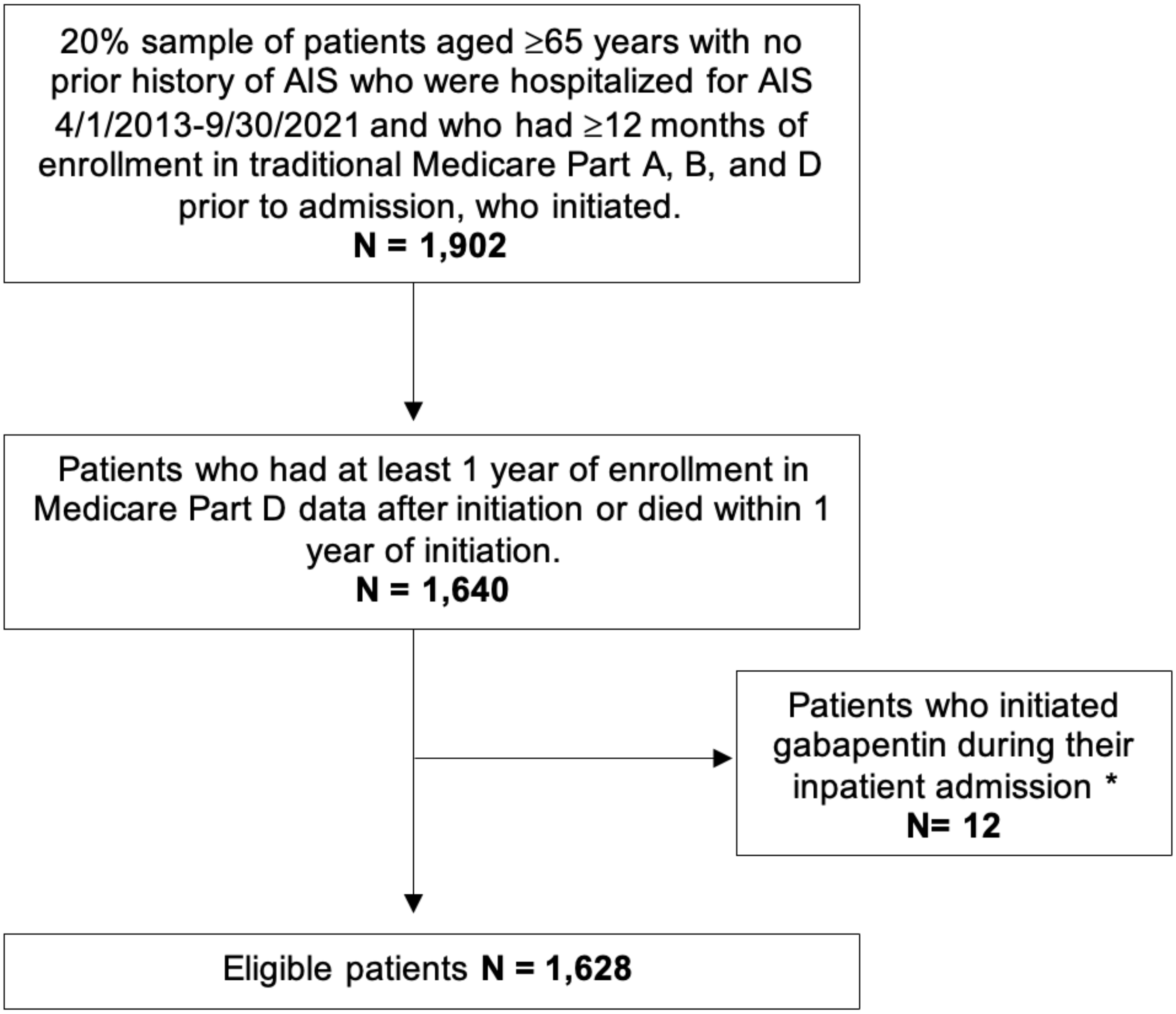


**Legend:** Medicare data files used: Medicare Provider Analysis and Review (MedPAR), Master Beneficiary Summary File (MBSF); Part D (Drug formulary data). Diagnosis for Acute Ischemic Stroke (AIS) based on ICD-9 codes 433, 434, and 436 and ICD-10 codes I63.x. Part A: Hospital Insurance, Part B: Medical Insurance; Part D: Drug coverage.

**Figure S2. Gabapentin treatment patterns were identified from the proportion of days covered (PDC) trajectories stratified by class**

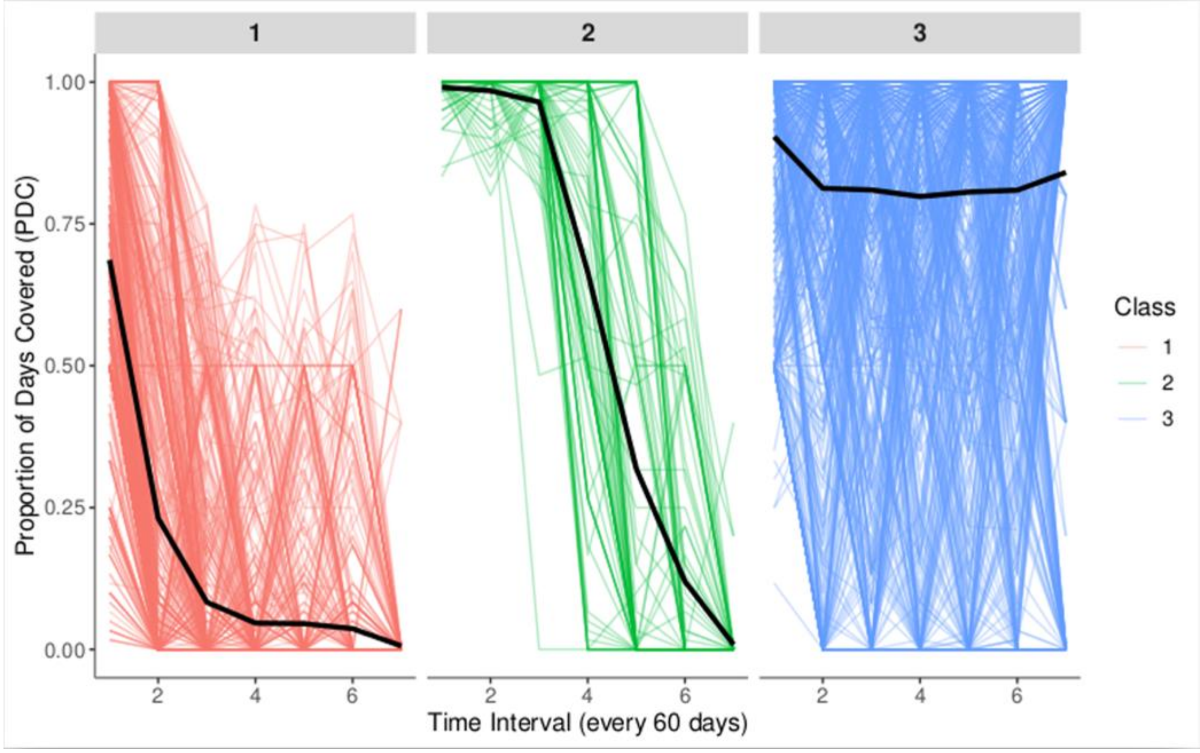


**Legend:** Solid lines denote the average PDC values in each class, where the minimum days of supply is smaller than 30 days, and 30 days is the median value.

### Supplemental Tables

**Table S1. STROBE Checklist**

| | Item # | Recommendation | Page # |
|---|---|---|---|
| **Title and abstract** | 1 | (*a*) Indicate the study's design with a commonly used term in the title or the abstract | 1 |
| | | (*b*) Provide in the abstract an informative and balanced summary of what was done and what was found | 4 |
| **Introduction** | | | |
| Background/ rationale | 2 | Explain the scientific background and rationale for the investigation being reported | 6 |
| Objectives | 3 | State specific objectives, including any prespecified hypotheses | 6-7 |
| **Methods** | | | |
| Study design | 4 | Present key elements of study design early in the paper | 7-8 |
| Setting | 5 | Describe the setting, locations, and relevant dates, including periods of recruitment, exposure, follow-up, and data collection | 7-8 |
| Participants | 6 | (*a*) *Cohort study*—Give the eligibility criteria, and the sources and methods of selection of participants. Describe methods of follow-up<br>*Case-control study*—Give the eligibility criteria, and the sources and methods of case ascertainment and control selection. Give the rationale for the choice of cases and controls<br>*Cross-sectional study*—Give the eligibility criteria, and the sources and methods of selection of participants | 7, Table S1 |
| | | (*b*) *Cohort study*—For matched studies, give matching criteria and number of exposed and unexposed<br>*Case-control study*—For matched studies, give matching criteria and the number of controls per case | |
| Variables | 7 | Clearly define all outcomes, exposures, predictors, potential confounders, and effect modifiers. Give diagnostic criteria, if applicable | 7-11 |
| Data sources/ measurement | 8* | For each variable of interest, give sources of data and details of methods of assessment (measurement). Describe comparability of assessment methods if there is more than one group | 8-9 |
| Bias | 9 | Describe any efforts to address potential sources of bias | |
| Study size | 10 | Explain how the study size was arrived at | 7-8, Table S1 |
| Quantitative variables | 11 | Explain how quantitative variables were handled in the analyses. If applicable, describe which groupings were chosen and why | 9, S3 |
| Statistical methods | 12 | (*a*) Describe all statistical methods, including those used to control for confounding | 10-11 |
| | | (*b*) Describe any methods used to examine subgroups and interactions | 10-11 |
| | | (*c*) Explain how missing data were addressed | 15 |
| | | (*d*) *Cohort study*—If applicable, explain how loss to follow-up was addressed | |

| | | | |
|---|---|---|---|
| | | *Case-control study*—If applicable, explain how matching of cases and controls was addressed<br>*Cross-sectional study*—If applicable, describe analytical methods taking account of sampling strategy | |
| | | (*e*) Describe any sensitivity analyses | 8, S3 |
| **Results** | | | |
| Participants | 13* | (a) Report numbers of individuals at each stage of study—eg numbers potentially eligible, examined for eligibility, confirmed eligible, included in the study, completing follow-up, and analysed | 11 |
| | | (b) Give reasons for non-participation at each stage | Table S1 |
| | | (c) Consider use of a flow diagram | Table S1 |
| Descriptive data | 14* | (a) Give characteristics of study participants (eg demographic, clinical, social) and information on exposures and potential confounders | 11-12 |
| | | (b) Indicate number of participants with missing data for each variable of interest | |
| | | (c) *Cohort study*—Summarize follow-up time (eg, average and total amount) | |
| Outcome data | 15* | *Cohort study*—Report numbers of outcome events or summary measures over time | |
| | | *Case-control study*—Report numbers in each exposure category, or summary measures of exposure | |
| | | *Cross-sectional study*—Report numbers of outcome events or summary measures | |
| Main results | 16 | (*a*) Give unadjusted estimates and, if applicable, confounder-adjusted estimates and their precision (e.g., 95% confidence interval). Make clear which confounders were adjusted for and why they were included | 11-12 |
| | | (*b*) Report category boundaries when continuous variables were categorized | |
| | | (*c*) If relevant, consider translating estimates of relative risk into absolute risk for a meaningful time period | |
| Other analyses | 17 | Report other analyses done—e.g. analyses of subgroups and interactions, and sensitivity analyses | |
| **Discussion** | | | |
| Key results | 18 | Summarize key results with reference to study objectives | 13-15 |
| Limitations | 19 | Discuss limitations of the study, taking into account sources of potential bias or imprecision. Discuss both direction and magnitude of any potential bias | 14-15 |
| Interpretation | 20 | Give a cautious overall interpretation of results considering objectives, limitations, multiplicity of analyses, results from similar studies, and other relevant evidence | 13-15 |
| Generalizability | 21 | Discuss the generalizability (external validity) of the study results | 13-15 |
| **Other information** | | | |
| Funding | 22 | Give the source of funding and the role of the funders for the present study and, if applicable, for the original study on which the present article is based | 16 |

**Table S2. Summary table of candidate models.**

| Model | # of classes | Covariates in class-membership model | Covariates in longitudinal model | Random effect | Log-likelihood | # of parameters | BIC | % Class 1 | % Class 2 | % Class 3 | % Class 4 |
|---|---|---|---|---|---|---|---|---|---|---|---|
| M1 | 1 | - | ~1+time+age+sex+seizure+depression+pain | time | -4505 | 10 | 9083 | 100% | | | |
| M1_ad | 1 | - | ~1+time+sex+seizure+pain | time | -4506 | 8 | 9070 | 100% | | | |
| M1_age | 1 | - | ~1+time+sex+seizure+depression+pain | time | -4505 | 9 | 9076 | 100% | | | |
| M1_asdp | 1 | - | ~1+time+sex | time | -4507 | 6 | 9058 | 100% | | | |
| M1_asex | 1 | - | ~1+time+seizure+depression+pain | time | -4505 | 8 | 9070 | 100% | | | |
| M1_dep | 1 | - | ~1+time+age+sex+seizure+pain | time | -4506 | 9 | 9078 | 100% | | | |
| M2_1 | 2 | ~1+age+sex+seizure | ~1+time+age+sex+seizure+depression+pain | time | -4484 | 16 | 9087 | 88.2% | 11.8% | | |
| M2_2 | 2 | ~1+age+sex+pain | ~1+time+age+sex+seizure+depression+pain | time | -4495 | 16 | 9109 | 7.0% | 93.0% | | |
| M2_3 | 2 | ~1+sex+sex+depression | ~1+time+age+sex+seizure+pain | time | -4497 | 15 | 9105 | 53.6% | 46.4% | | |
| M2_4 | 2 | ~1+age+sex | ~1+time+age+sex+seizure+pain | time | -4498 | 14 | 9099 | 47.8% | 52.2% | | |
| M2_5 | 2 | ~1+sex | ~1+time+age+sex+seizure+depression+pain | time | -4486 | 14 | 9075 | 94.8% | 5.2% | | |
| M2_6 | 2 | ~1+age | ~1+time+age+sex+seizure+depression+pain | time | -4517 | 14 | 9137 | 49.9% | 50.1% | | |
| M2_7 | 2 | ~1+age | ~1+time+sex+seizure+depression+pain | time | -4498 | 13 | 9091 | 12.5% | 87.5% | | |
| M2_8 | 2 | ~1 | ~1+time+age+sex+seizure+depression+pain | time | -4486 | 13 | 9068 | 5.3% | 94.7% | | |
| M2_9 | 2 | ~1+age | ~1+time+sex+seizure+pain | time | -4499 | 12 | 9087 | 11.4% | 88.6% | | |
| M2_10 | 2 | ~1 | ~1+time+sex+seizure+pain | time | -4488 | 11 | 9058 | 5.3% | 94.7% | | |
| M2_11 | 2 | ~1+seizure | ~1+time+sex+seizure+pain | time | -4488 | 12 | 9065 | 5.3% | 94.7% | | |
| M2_12 | 2 | ~1+age+sex | ~1+time+seizure+depression+pain | time | -4498 | 13 | 9092 | 9.0% | 91.0% | | |
| M2_13 | 2 | ~1+age+sex | ~1+time+seizure | time | -4500 | 11 | 9081 | 8.5% | 91.5% | | |
| M3_5 | 3 | ~ 1+sex | ~1+time+age+sex+seizure+depression+pain | time | -4481 | 18 | 9095 | 16.8% | 73.5% | 9.8% | |
| **M3_8** | **3** | **~1** | **~1+time+age+sex+seizure+depression+pain** | **time** | **-4468** | **16** | **9054** | **42.5%** | **5.9%** | **51.6%** | |
| M3_10 | 3 | ~1 | ~1+time+sex+seizure+pain | time | -4476 | 14 | 9055 | 48.7% | 44.9% | 6.4% | |
| M3_11 | 3 | ~1+seizure | ~1+time+sex+seizure+pain | time | -4481 | 16 | 9080 | 49.6% | 5.4% | 45.0% | |
| M3_13 | 3 | ~1+age+sex | ~1+time+seizure | time | -4488 | 16 | 9093 | 19.5% | 37.3% | 43.2% | |
| M4_8 | 4 | ~1 | ~1+time+age+sex+seizure+depression+pain | time | -4459 | 19 | 9058 | 47.1% | 5.8% | 4.4% | 42.7% |
| M4_10 | 4 | ~1 | ~1+time+sex+seizure+pain | time | -4466 | 17 | 9058 | 7.4% | 5.7% | 43.1% | 43.9% |

**Table S3. Latent class mixed model results**

| Fixed effects in the class-membership model (class 3 is the reference) | | | | |
|---|---|---|---|---|
| | coefficient | standard error | test statistic | p-value |
| Intercept class 1 | -0.44 | 0.12 | -3.72 | 0.00 |
| Intercept class 2 | -2.21 | 0.34 | -6.48 | 0.00 |
| Fixed effects in the longitudinal model | | | | |
| | coefficient | standard error | test statistic | p-value |
| Intercept class 1 (not estimated) | - | - | - | - |
| Intercept class 2 | -5.97 | 2.39 | -2.50 | 0.01 |
| Intercept class 3 | 0.91 | 0.36 | 2.52 | 0.01 |
| Time class 1 | 1.94 | 0.29 | 6.71 | 0.00 |
| Time class 2 | 2.03 | 0.47 | 4.31 | 0.00 |
| Time class 3 | -0.02 | 0.02 | -0.95 | 0.34 |
| Age | -0.06 | 0.04 | -1.37 | 0.17 |
| Female | -0.11 | 0.11 | -0.99 | 0.32 |
| Baseline Seizure | 1.42 | 0.50 | 2.85 | 0.00 |
| Baseline Depression | -0.27 | 0.17 | -1.58 | 0.11 |
| Baseline Pain | -0.09 | 0.53 | -0.17 | 0.87 |

### R Codes

```
#---- PDC trajectories ----
#---- PDC: global functions ----
# PDC
Overlap_loop = function(data){

  data$EndDate_a = data$DateService_a = rep(as.Date(NA), nrow(data))
  data$EndDate_a[1] = data$EndDate[1]
  data$DateService_a[1] = data$DateService[1]

  if (nrow(data) > 1) {

  for(i in 2:nrow(data)){

    data$EndDate_a[i] = data$EndDate[i]
    data$DateService_a[i] = data$DateService[i]

    if (data$EndDate_a[i-1]>=data$DateService_a[i])
    {
      data$DateService_a[i] = data$EndDate_a[i-1]+1
      data$EndDate_a[i] = data$DateService_a[i]+data$Duration[i]-1
    }
  }
  }
  return(data)
}

# revised: stretch the date_a and count missing days (if we delay the overlapped prescription)
# any refill or renewal >=90 days, treat patient as non-persistence
One_QteTS_a = function(data, slice_window=60, obs_window=365*1, non_pers){

  if(nrow(data) != 1) {

    #detect non-persistence
    data = data %>% mutate(TI = DateService_a-lag(DateService_a, default = first(DateService_a)),
                       TI2 = ifelse(Renouvel_adj=="N", NA, TI),
                       gap = TI2 - lag(Duration, default = first(Duration)))
           }
  }
  # Non-persistency is not a censoring factor
  if(sum(data$death_cens_ind) >= 1){
    QteTS = data.frame(date=seq.Date(from=min(data$DateService_a),
                                 to=max(data$EndDate_a),
                                 by="days"))
  } else {
    QteTS = data.frame(date=seq.Date(from=min(data$DateService_a),
```

```
                                to=min(data$DateService_a)+obs_window-1,
                                by="days"))
  }

  TS = data %>% mutate(date=map2(DateService_a, EndDate_a, seq, "day")) %>% unnest(date)
  QteTS = QteTS %>%
    left_join(TS, by="date") %>%
    mutate(flag = na.locf(Renouvel_adj, fromLast = TRUE, na.rm=FALSE))

  #gap_rmN indicates: gap days indicator (0: gap days)

  QteTS$gap_rmN = ifelse(is.na(QteTS$Renouvel_adj)&QteTS$flag=="R", 0, 1)
  QteTS$gap_rmN = ifelse(is.na(QteTS$flag),0,QteTS$gap_rmN)

  # count 0s every @slice_window days

  QteTS2 = QteTS %>%
    #filter(!is.na(Duration)|flag!="N") %>%
    mutate(day = seq(1, n(),1), interval_id = (day-1)%/%slice_window+1) %>%
    group_by(interval_id) %>%
    summarise(zero_cnt_rmN = sum(gap_rmN==0),
          day_length = n(),.groups = 'drop',
          LCMA1 = (day_length-zero_cnt_rmN)/day_length)

  # overall PDC & time to non-adherence

  QteTS2$cum_PDC = sum(QteTS2$day_length-QteTS2$zero_cnt_rmN)/sum(QteTS2$day_length)
  return(QteTS2)
}

# get first claim date, number of refills, claim number, and gap between refills
rx_pdc <- df_rx_sum_admit_cen %>%
  group_by(BENE_ID) %>%
  arrange(DateService, EndDate, .by_group = T) %>%
  # edit here
  #filter(n()>1) %>% # at least one refill to calculate PDC
  mutate(MinDate = min(DateService,na.rm=T),
       NumberRefills = n(),
       PrescriptionNumber = row_number(),
       Gap = as.numeric(DateService - lag(EndDate))) %>%
  mutate(Refill = ifelse(DateService == MinDate, "N", "R"))


# adjusted prescription start date and end date
# censor non-persistence
rx_pdc_adj = rx_pdc %>%
  group_by(BENE_ID) %>%
  arrange(DateService, EndDate, .by_group = T) %>%
```

```
  #filter(BENE_ID=="zzzzzzewPPepPpp") %>%
  group_modify(~Overlap_loop(.)) %>%
  #mutate(EndDate_a = if_else(row_number()==n(), DateService_a, EndDate_a)) %>%
  mutate(cumInterval_persist=lead(DateService_a,1)-EndDate_a,
       FU_cens_date = as.Date(FU_cens_date)) %>%
  # adjust refills within obserational windows
  filter(DateService_a <= FU_cens_date) %>%
  mutate(EndDate_a = if_else(row_number()==n() & (EndDate_a > FU_cens_date), FU_cens_date, EndDate_a))
%>%
  ungroup()

# adherence trajectories
rx_pdc_traj = rx_pdc_adj %>% rename(Renouvel_adj = Refill) %>%
  dplyr::select(BENE_ID, DateService_a, EndDate_a, Renouvel_adj, Duration, FU_cens_date) %>%
  left_join(df_rx_30 %>% select(BENE_ID, initiation_date, BENE_DEATH_DT) %>% distinct()) %>%
  mutate(death_cens_ind = if_else(FU_cens_date==BENE_DEATH_DT, 1, 0)) %>%
  mutate(death_cens_ind = if_else(is.na(death_cens_ind),0,death_cens_ind)) %>%
  group_by(BENE_ID) %>%
  arrange(DateService_a, EndDate_a, .by_group = T) %>%
  group_modify(~One_QteTS_a(.,slice_window=60, obs_window=obs_days, non_pers = nonpers_day))

#latent class mixed model
# df_rx_demo contains demographic variables
rx_pdc_traj_cov = rx_pdc_traj %>% left_join(df_rx_demo, by="BENE_ID") %>% left_join(unique_ids, by =
"BENE_ID") %>% droplevels()
rx_pdc_traj_cov$pdc_binary = ifelse(rx_pdc_traj_cov$LCMA1<0.8, 1, 0)

m3 = lcmm(pdc_binary ~ interval_id + age + sex + seizure + depression + pain,
          random = ~ interval_id,
          subject = "ID",
          mixture = ~ interval_id,
          classmb = ~ 1,
          ng = 3, B = random(m0),
          data = rx_pdc_traj_cov, link = "thresholds")
```
